# Automated processing of thermal imaging to detect COVID-19

**DOI:** 10.1101/2020.12.22.20248691

**Authors:** Rafael Y. Brzezinski, Neta Rabin, Nir Lewis, Racheli Peled, Ariel Kerpel, Avishai M. Tsur, Omer Gendelman, Nili Naftali-Shani, Irina Gringauz, Howard Amital, Avshalom Leibowitz, Haim Mayan, Ilan Ben-Zvi, Eyal Heler, Liran Shechtman, Ori Rogovski, Shani Shenhar-Tsarfaty, Eli Konen, Edith M. Marom, Avinoah Ironi, Galia Rahav, Yair Zimmer, Ehud Grossman, Zehava Ovadia-Blechman, Jonathan Leor, Oshrit Hoffer

## Abstract

Rapid and sensitive screening tools for SARS-CoV-2 infection are essential to limit the spread of COVID-19 and to properly allocate national resources. Here, we developed a new point-of-care, non-contact thermal imaging tool to detect COVID-19, based on image-processing algorithms and machine learning analysis. We captured thermal images of the back of individuals with and without COVID-19 using a portable thermal camera that connects directly to smartphones. Our novel image processing algorithms automatically extracted multiple texture and shape features of the thermal images and achieved an area under the curve (AUC) of 0.85 in detecting COVID-19 with up to 92% sensitivity. Thermal imaging scores were inversely correlated with clinical variables associated with COVID-19 disease progression. We show, for the first time, that a hand-held thermal imaging device can be used to detect COVID-19. Non-invasive thermal imaging could be used to screen for COVID-19 in out-of-hospital settings, especially in low-income regions with limited imaging resources.

**HIGHLIGHTS:**

- Automated processing of thermal images of the back can be used to detect COVID-19 with up to 92% sensitivity.
- The extracted texture features of the thermal image are associated with COVID-19 disease progression and lung injury.
- A portable thermal camera that connects directly to smartphones can be used to detect COVID-19.
- Non-invasive thermal imaging could be used to screen for COVID-19 in out-of-hospital settings and regions with limited imaging resources.

**GRAPHICAL ABSTRACT:** 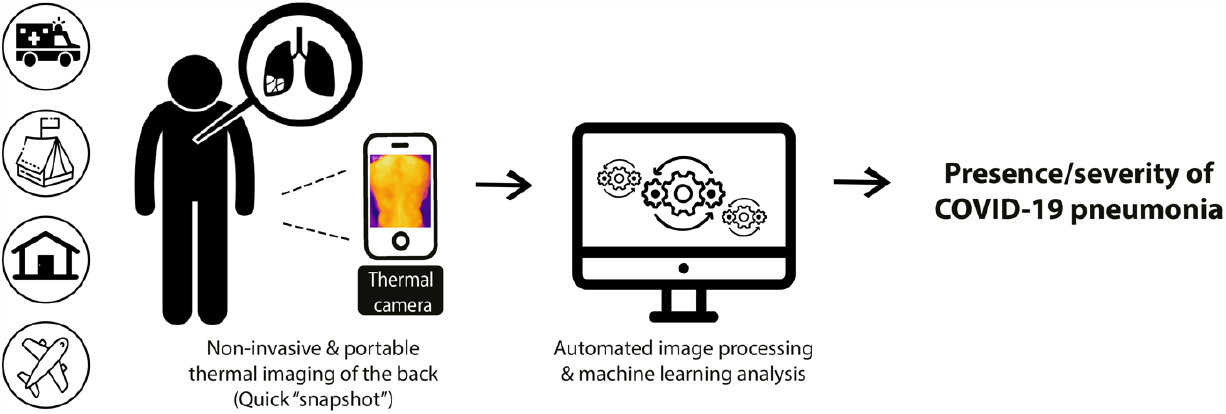

## 1. INTRODUCTION

The coronavirus disease 2019 (COVID-19) pandemic has imposed an immense burden on out-of-hospital health care services and community-based testing sites worldwide^1,2^. Immediate and sensitive screening tools for severe acute respiratory syndrome coronavirus 2 (SARS-CoV-2) infection are essential in order to limit the spread of the disease and to properly allocate national resources. The common and standard diagnosis of SARS-CoV-2 infection is based on a virus-specific quantitative real-time PCR (qRT-PCR). The fastest test, however, can take up to several hours to complete, depending on available resources and disease burden in the region^3^. Furthermore, a significant amount of skilled labor is required at the different stages of qRT-PCR testing (sampling, preparation, and analysis)^3^.

Infrared thermography scanners have been widely used as a screening tool during past infectious disease epidemics, including COVID-19^4,5^. This technique is based on the assessment of absolute body temperature in order to screen individuals with fever and lack specificity for COVID-19^5^.

Non-invasive thermal imaging of organ-specific diseases has been presented as a new tool to detect inflammation and vascular dysfunction^6–8^. We propose that advanced processing of thermal images, combined with machine learning analysis, could serve as an innovative diagnostic tool for detecting COVID-19 and its associated microvascular injury.

Here, we developed automated image processing algorithms for thermal images of the back, captured by a portable thermal camera that connects directly to smartphones. We aimed to determine the ability of this novel technique to detect COVID-19-associated lung injury. Our automated thermal imaging tool is relatively cheap, easy to use, and delivers immediate test results. It is therefore especially applicable for out-of-hospital screening by first responders (paramedics, nurses, medical technicians etc.), home care facilities, military bases, and regions with limited diagnostic resources.

## 2. RESULTS

### 2.1 Study cohort

The final study cohort included 101 individuals who were prospectively enrolled from two medical centers in Israel (**Table 1**). The mean age was 56 ± 17 years, and 80 participants (79%) were men. A total of 62 (61%) individuals were COVID-19 positive, of whom 40 (85%) had some form of lung injury, defined as any clinical or radiological evidence of pneumonia or acute respiratory distress^9^, including hypoxemia, dyspnea, respiratory rate >20, shortness of breath, severe cough, and/or the presence of ground glass appearance, consolidation, or linear opacities on chest X-ray (CXR).

**Table 1:** Patient characteristics.

| Characteristic | COVID-19 negative | COVID-19 positive | p |
| --- | --- | --- | --- |
|  | n = 39 | n = 62 |  |
| Age, years | 48.8 ± 20 | 60.6 ± 13 | 0.01 |
| Sex (male), n (%) | 31 (80%) | 49 (79%) | 0.9 |
| Lung injury, n (%) | 7 (18%) | 40 (65%) | 0.01 |
| Body temperature, °C | 37.1 ± 0.5 | 36.9 ± 0.6 | 0.3 |
| SPO<sub>2</sub> room air, % | 95.7 ± 2.9 | 94.4 ± 4.2 | 0.3 |
| Hypertension, n (%) | 10 (26%) | 24 (39%) | 0.2 |
| Dyslipidemia, n (%) | 10 (26%) | 28 (45%) | 0.05 |
| Diabetes, n (%) | 6 (15%) | 15 (24%) | 0.3 |
| CRP, mg/L | 14.1 [2.0, 96.5] | 47.9 [11.8, 120.4] | 0.5 |
| D-dimer, mg/ml | 401 [187, 792] | 503 [174.1, 745] | 0.4 |
| WBC, 10<sup>9</sup>/L | 8.7 [7.1, 10.9] | 6.3 [5, 7.9] | 0.06 |
| Cardiac troponin-I, ng/L | 3.3 [2.7, 13.3] | 6.2 [4, 12.1] | 0.7 |
| Hemoglobin, g/dL | 12.2 ± 2.5 | 13.6 ± 2.3 | 0.03 |

*Values are presented as mean ± SD, or median [interquartile range] for irregular distributed parameters. Blood test results were taken from the day of imaging (n = 78). CRP: C-reactive protein; WBC- white blood count; SPO<sub>2</sub>- Saturation of Peripheral Oxygen. P values by two-tailed unpaired t-test, the non-parametric Mann-Whitney test, and a Chi-square test.*

COVID-19 positive patients were older than COVID-19 negative individuals and had a higher prevalence of lung injury (**Table 1**, 65% vs. 18%, p < 0.01). Absolute body temperature was similar in both groups, as was as the prevalence of hypertension, dyslipidemia and diabetes (**Table 1**). Overall, the majority of COVID-19 patients included in our study were ultimately discharged with a favorable prognosis. Only three patients required mechanical ventilation during follow-up; two eventually died (one with and one without COVID-19).

### 2.2 Thermal image processing detected COVID-19 and lung injury status

We captured thermal images of the participants’ exposed upper backs (over the lungs) using a portable thermal camera that connects directly to smartphones. All images of the final study cohort (n = 101) were analyzed by investigators who were blinded to the patients’’ clinical data. Our novel thermal image processing algorithms extracted over 100 texture and shape features of temperature distribution across the skin. Two key parameters were chosen for downstream analysis: the fractal dimension of the gradient (termed FD) and a new parameter we called the “sum of extrema” in the image (termed SX-details in Methods) (**Figure 1**). Both FD and SX scores were significantly lower in COVID-19 positive individuals compared to the rest of the cohort (**Figure 2**).

**Figure 1:**
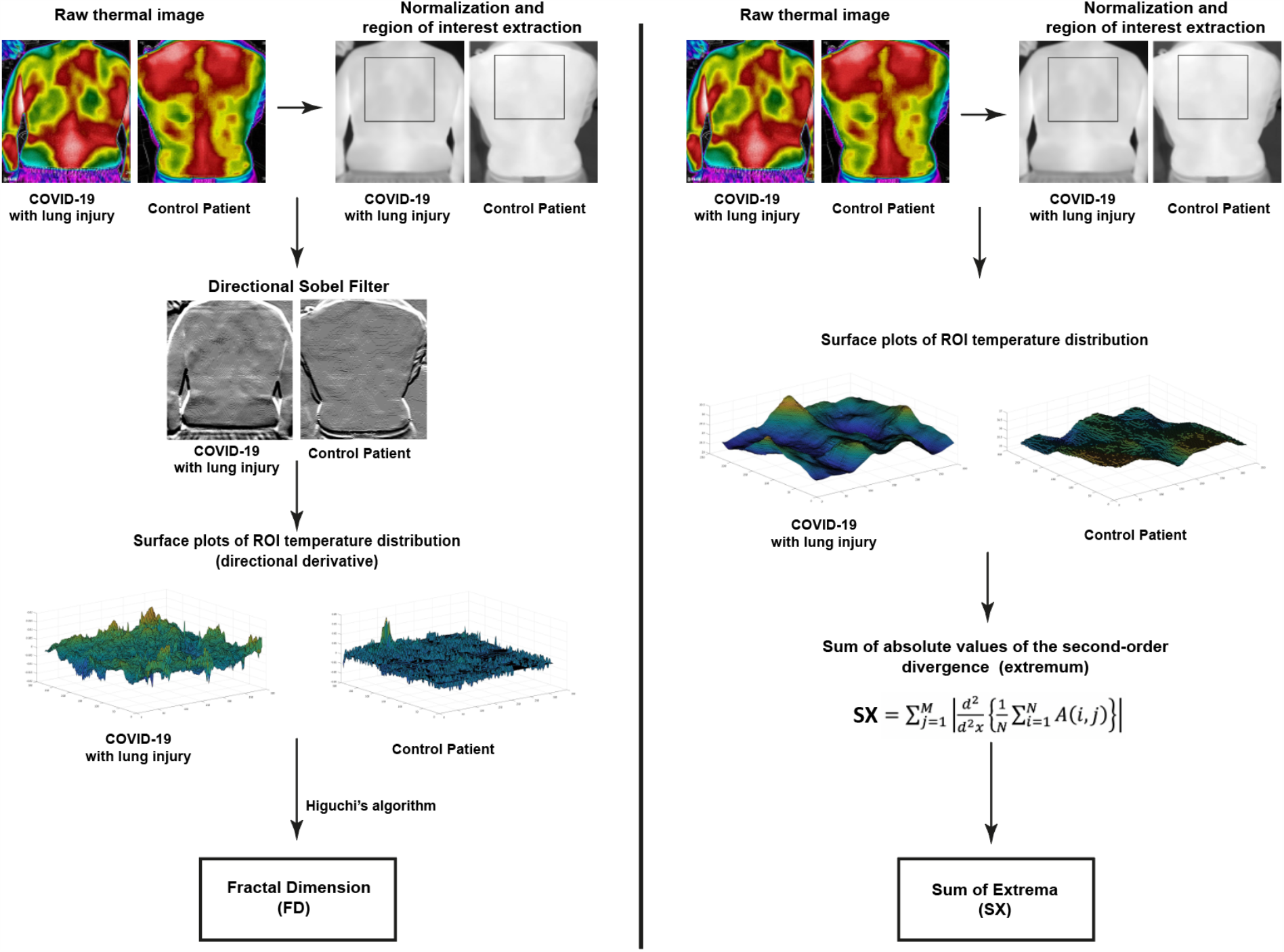
Thermal image processing. The raw thermal images are displayed in HC-Rainbow scale. Displayed are representative images of the different stages of our image processing algorithms for the fractal dimension (FD) of the gradient (left) and the “sum of extrema” (SX) in the image (right). For this display the gradient range [-8,8] was normalized to [0,255]. All gradient values outside of this range were shifted to its edges before normalization. Full details on image processing algorithms are provided in the Methods section. *ROI - region of interest*.

**Figure 2:**
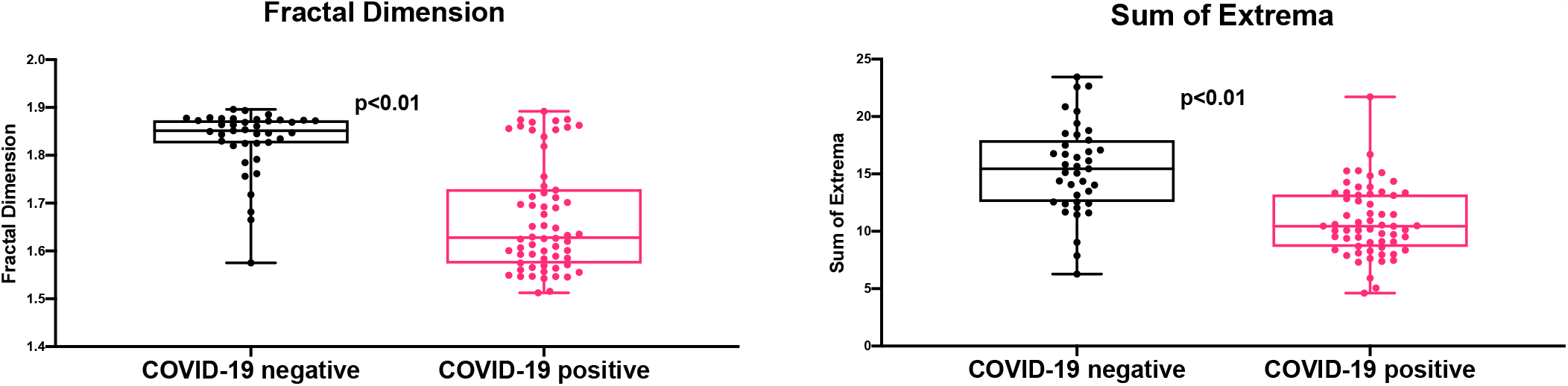
Thermal imaging scores according to COVID-19 disease status. Box plots with individual values of fractal dimension (FD) (left side) and sum of extrema (SX) (right side) scores. Patients who were COVID-19 positive had lower thermal scores compared to COVID-19 negative individuals. P value by unpaired T-test.

Next, we evaluated the ability of our thermal scores to detect COVID-19 disease status and/or the presence of lung injury. Sensitivity, specificity, and area under the curve (AUC) were calculated for both FD and SX scores. The receiver operating characteristic (ROC) curves (**Figure 3**) indicated that both FD and SX scores were significantly associated with the diagnosis of COVID-19, with an AUC of 0.85 (95% CI: 0.78, 0.93; p < 0.01) and 0.82 (95% CI: 0.73, 0.91; p < 0.01) respectively. A combined cutoff of either a low FD score (≤ 1.82) and/or low SX score (≤ 13.5) demonstrated a 92% sensitivity and 62% specificity in detecting COVID-19 status (regardless of lung injury) (**Figure 3**). Thus, the two thermal scores FD and SX were able to identify COVID-19 in suspected individuals.

**Figure 3:**
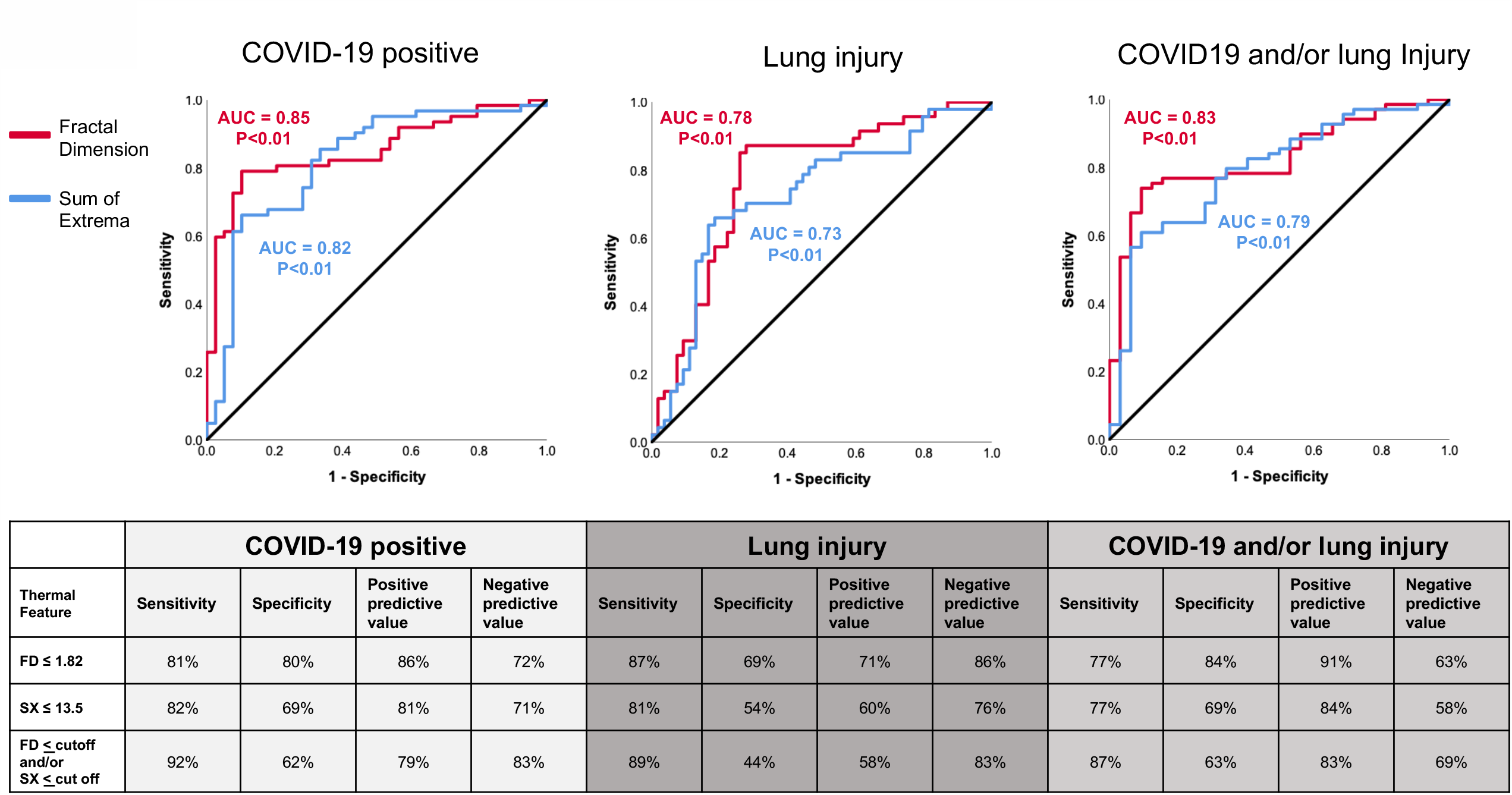
The diagnostic yields of thermal imaging scores in detecting COVID-19 and lung injury. Receiver operating characteristic (ROC) curves are shown (top panel) for the three different clinical classifications. Sensitivity and specificity values were calculated for select cutoffs (bottom panel).

FD and SX scores were also significantly associated with lung injury, with an AUC of 0.78 (95% CI: 0.69, 0.88, p < 0.01) and 0.73 (95% CI: 0.63, 0.83, p < 0.01) (**Figure 3**). The combined cutoff of either a low FD score and/or low SX score demonstrated a high sensitivity of 89% but low a specificity of 44% in detecting lung injury (regardless of COVID-19 status) (**Figure 3**). FD outperformed SX in detecting the composite diagnosis of COVID-19 and/or lung injury, with an AUC of 0.83 (95% CI: 0.75, 0.91, p<0.01) vs. 0.79 (95% CI: 0.70, 0.88, p<0.01).

To determine the effect of the anatomical location of the selected region of interest (ROI) on diagnostic yields, we ran our algorithms on a different ROI: the lower back region, below the lungs (**Supp Fig**. 1). Both FD and SX scores demonstrated similar diagnostic yields for detecting COVID-19 status compared with the upper back ROI selection model: AUC = 0.85 (95% CI: 0.76, 0.93, p < 0.01) for FD and 0.79 (95% CI: 0.7, 0.88, p < 0.01) for SX. Thus, it seems that the differences in temperature distribution associated with COVID-19 are not restricted to the skin covering the lungs, but rather reflect a systemic pattern present across the entire torso.

### 2.3 The correlation between thermal imaging scores and clinical variables

Next, we evaluated the correlation between our thermal scores and selected clinical variables that were shown to predict poor prognosis in COVID-19 patients^10,11^. Patients with saturation of peripheral oxygen (SPO_2_) ≤ 93% on the day of imaging had lower FD and SX scores (**Figure 4A**). Moreover, even patients who had normal SPO_2_ values on the day of imaging but developed hypoxia (≤ 93%) during follow-up, had lower FD and SX scores on thermal imaging (**Figure 4A**), suggesting a possible early indication of clinical deterioration seen by thermal imaging. In line with these observations, patients who required non-invasive oxygen support such as nasal cannula or face masks had lower FD scores than patients who did not need oxygen therapy: 1.67± 0.07 vs. 1.74± 0.13, p < 0.01. Overall, low blood oxygen saturation was associated with low FD and SX thermal scores.

**Figure 4:**
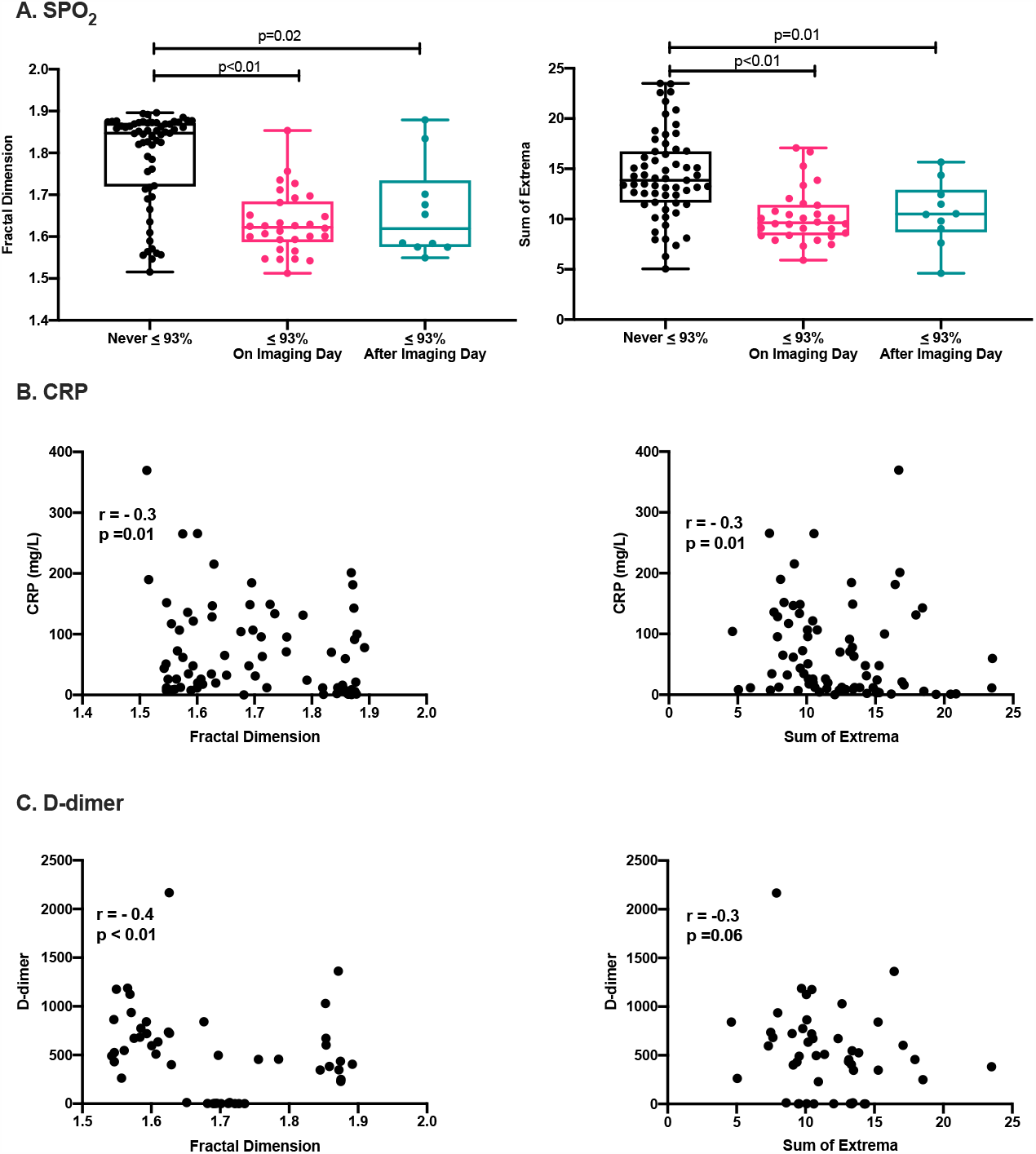
Thermal imaging scores and clinical measurements. **A**. Box plots with individual measurements of the fractal dimension (FD) (left) and sum of extrema (SX) (right). P value by one-way ANOVA with Tukey’s test for multiple comparisons. **B-C**. The correlation between the thermal FD and SX scores and C-reactive protein (CRP) (B) and D-Dimer (C) measured on the imaging day. P value by Spearman’s correlation test.

C-reactive protein (CRP) and D-dimer levels are associated with COVID-19 severity and prognosis^10,11^. Significantly, the thermal FD and SX scores were inversely correlated with CRP and D-dimer levels measured on the imaging day (**Figure 4B+C**). We did not find a correlation between FD and SX with cardiac troponin-I concentrations: r = 0.04, p= 0.78, and r = −0.22, p = 0.11. Taken together, the thermal FD and SX scores were inversely correlated with clinical variables that reflect COVID-19 disease progression.

### 2.4 Thermal imaging scores did not correlate with the findings on chest X-ray

To compare our thermal imaging technique to conventional CXR, we used the radiographic assessment of lung edema (RALE) score^12^. We compared thermal imaging scores with RALE scores for all patients who had a CXR within 6 hours of thermal imaging (n = 30). The CXRs were manually reviewed by a certified radiologist who was blinded to the patients’’ clinical statuses and thermal imaging scores.

First, in line with a previous report^12^, RALE scores were not different between patients with and without COVID-19, and did not provide diagnostic yields (**Figure 5A**). As expected, patients with lung injury had higher RALE scores than control patients (**Figure 5A**). However, patients with a pathological CXR (RALE score>0) and patients with a normal CXR (RALE score=0) had similar FD and SX scores (**Figure 5B**). Both thermal FD and SX scores were not correlated with RALE scores across the entire cohort (**Figure 5C**). Thus, thermal imaging scores did not correlate with the findings on CXR.

**Figure 5:**
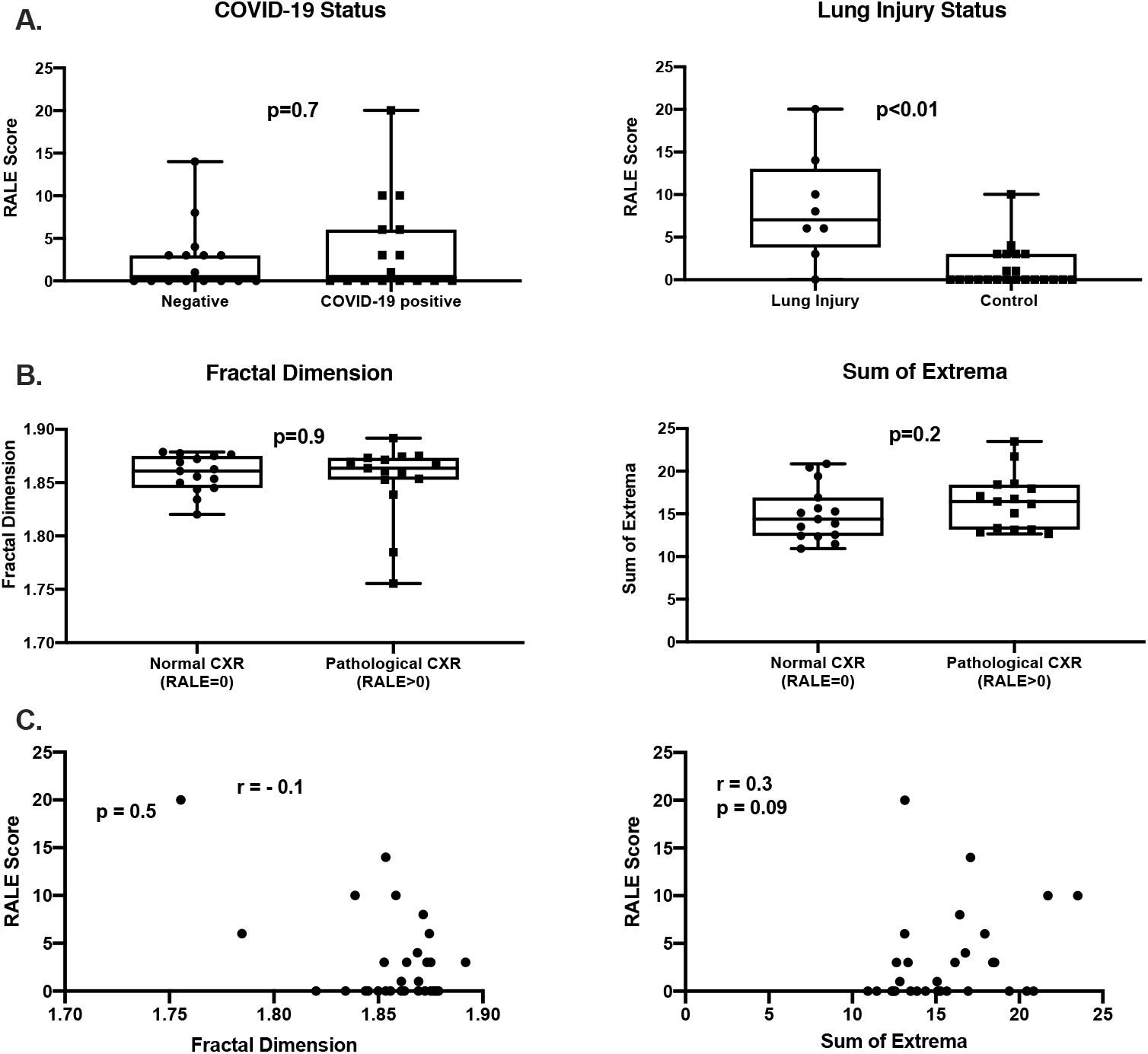
The correlation between thermal imaging and chest X-ray. **A**. Box plots of RALE score according to COVID-19 and lung injury status. P value by Mann-Whitney test. **B**. Box plots of the fractal dimension (FD) (left) and sum of extrema (SX) (right) thermal scores according to categorized RALE score. P value by Mann-Whitney test. **C**. RALE scores were not significantly correlated with. P value by Spearman’s correlation test. *CXR-Chest X-ray*; *RALE - radiographic assessment of lung edema*.

### 2.5 Prediction of COVID-19 status based on thermal features and machine learning techniques

Finally, to validate our ability to detect COVID-19 patients, the dataset was split into two groups: a training set (81) and a validation set (20). The predicted label was COVID-19 status. Three machine learning classification models were tested on the training set in a leave-one-out-cross-validation manner: logistic regression, k-nearest neighbors (k-nn) and kernel-support vector machine (SVM). The kernel-SVM model demonstrated up to 88% sensitivity, while the logistic regression model demonstrated the highest specificity of 84% on the training set and 100% on the validation set (**Supp Table 1**). A 3-dimensional plot of the logistic regression model is shown in **Figure 6**. The addition of age and sex to the models did not improve accuracy, while the specificity of the k-nn model was degraded (**Supp Table 1**).

**Figure 6:**
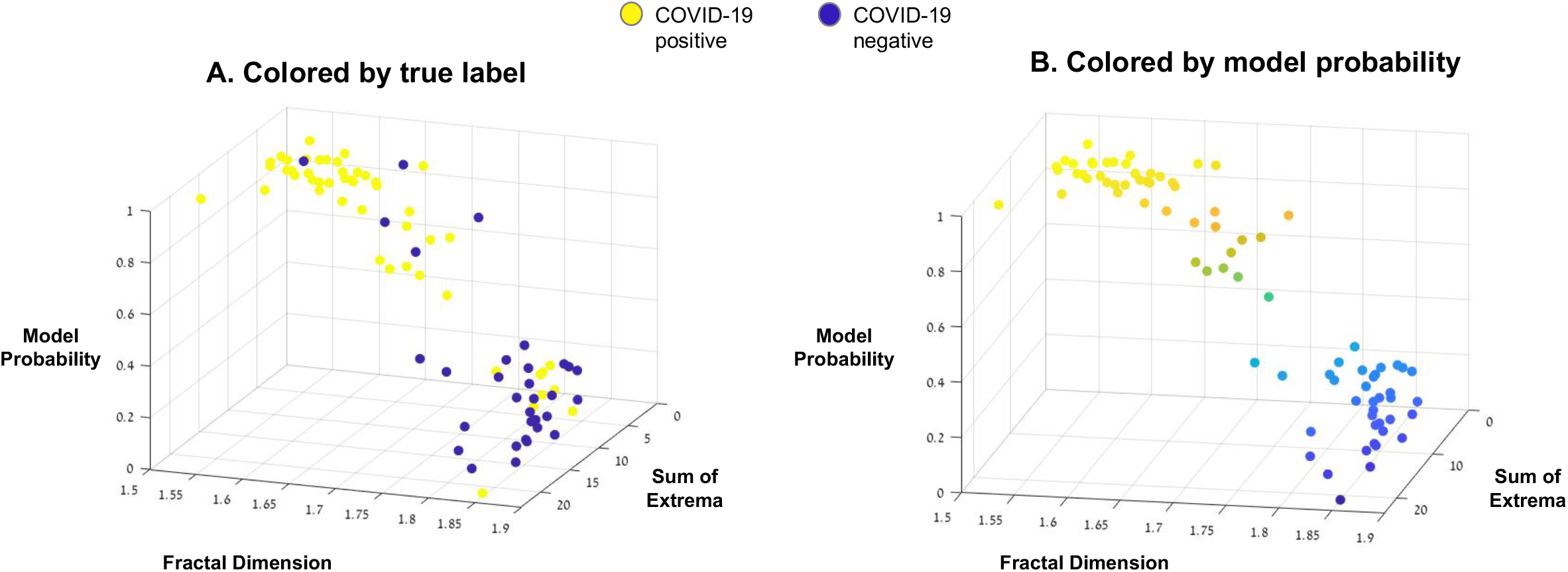
Logistic regression model to predict COVID-19 status. A 3-dimensional plot of the logistic regression model on the training data (N = 81) is shown, colored by the true label of the patient (A) and the model probability (B) by color spectrum from yellow (COVID-19 positive) to purple (COVID-19 negative). Newly arrived patients from the validation set were classified accordingly (Supp Table 1).

Overall, machine learning based analysis of the two thermal FD and SX scores was able to classify individuals according to their COVID-19 status, independent of any other clinical variables.

## 3. DISCUSSION

We suggest, for the first time, that a hand-held thermal imaging device that connects directly to smartphones can detect individuals infected with SARS-CoV-2 (**Figure 7**). Automated thermal image processing of the back yielded two risk scores that demonstrated up to 92% sensitivity in detecting COVID-19.

**Figure 7:**
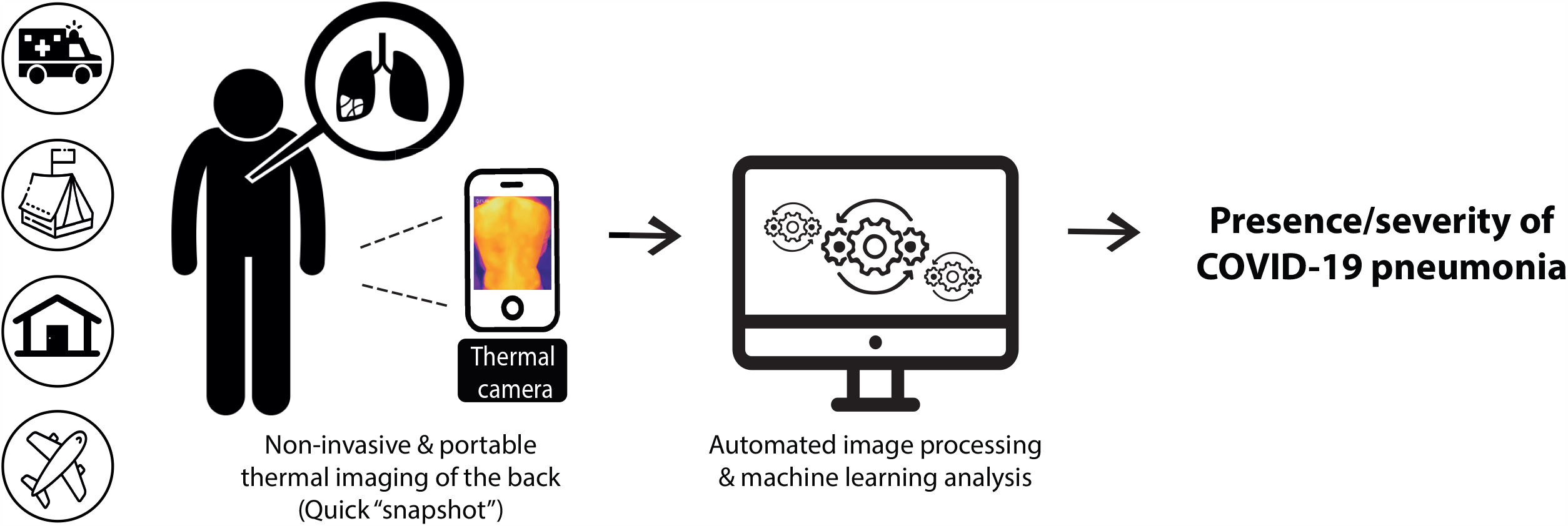
Graphical Abstract. A schematic illustration of the research design and potential impact.

Portable thermal imaging has several key advantages over currently available diagnostic alternatives for COVID-19 patients. Primarily, this method is well suited for out-of-hospital settings and low-income regions that have limited access to more advanced imaging modalities such as ultrasound, CT, and MRI. Our thermal imaging technique does not require prolonged training or expertise and is compatible with low-cost thermal cameras, such as the one used in this study (FLIR ONE). Portable thermal imaging could be used to screen suspected individuals for COVID-19 and/or associated pneumonia in out-of-hospital and quarantine settings, as well as in nursing homes, in the field and in home-hospitalization cases. Non-contact thermal imaging does not expose patients to ionizing radiation, does not require disposable reagents, and could become a first-line diagnostic tool for COVID-19, thus aiding in early triage and resource allocation in low-income regions.

Current efforts to use infrared thermography in the fight against the COVID-19 pandemic have focused primarily on measuring absolute body temperature and screening for individuals with fever in crowded settings^4,5^. However, these attempts have demonstrated limited effectiveness and were not shown to have a substantive impact in blocking the spread of COVID-19^4,5^. Our innovative approach to extract advanced texture and shape features from the thermal image could help improve these screening tools and aid in the diagnosis of other end-organ damage associated with COVID-19, such as cardiac dysfunction and acute kidney injury^13,14^. The fact that absolute body temperature did not differ between COVID-19 positive and negative patients in our cohort supports our rationale to focus on temperature distribution across organs of interest, as opposed to standard temperature assessment.

The physiological basis of our findings is not entirely clear. We suggest that the differences in skin temperature distribution across the back, as measured by our advanced texture features FD and SX, may reflect microvascular dysfunction caused by COVID-19^15,16^. The fact that both thermal features were inversely correlated with CRP and D-dimer concentrations supports the concept that these thermal scores reflect elevated systemic inflammation in infected individuals, and especially so in patients with lung injury. Microvascular dysfunction has been associated with abnormal temperature distribution across the skin^17,18^. Microvascular and endothelial dysfunction are also characteristic of COVID-19^15,16^. Thus, thermal imaging might detect micro-angiopathies and endothelial dysfunction in COVID-19 patients and could possibly improve risk stratification of infected individuals.

Our study has several limitations. First, we used relatively small sample sizes. Second, the cutoffs used for our thermal scores to detect COVID-19 and lung injury were optimized for this specific study cohort, and need to be adjusted in future multi-center trials with larger study populations that are more gender and ethnically diverse. Future large scales efforts might also improve the relatively low specificity levels of our thermal scores in detecting COVID-19 and lung injury. Finally, data on smoking history and BMI were unavailable for the majority of the cohort due to missing variables in electronic health records. Smoking status and chronic lung injury may have an effect on thermal imaging scores and this aspect will need to be addressed in future studies.

In summary, we have developed here a non-contact portable thermal imaging tool to detect COVID-19 and lung injury (**Figure 7**). This technique could facilitate the screening of large numbers of people in order to detect infected individuals in real time, limit the spread of COVID-19, and aid in the allocation of resources throughout the healthcare system.

## 4. METHODS

### 4.1 Study cohort

We recruited individuals with and without COVID-19 from two medical centers in Israel: the Sheba Medical Center, Ramat Gan, and the Tel Aviv Sourasky Medical Center, Tel Aviv. Healthy individuals were also recruited from the Afeka Tel Aviv College of Engineering, Tel Aviv. The study was approved by the institutional review boards of each participating medical center or academic institution, and participants provided verbal (recording) or written informed consent prior to their participation in the study.

The research design and patient selection process are presented in **Supp Fig. 2**. Briefly, we recruited individuals from three settings: 1) emergency room (ER) patients (n = 37); 2) patients hospitalized at a designated COVID-19 ward or a regular internal medicine ward (for COVID-19 negative patients) (n = 55); and 3) apparently healthy individuals from all three institutions (n = 14).

We included adults (aged 18-90 years) with suspected pneumonia and/or COVID-19 who had undergone a CXR or CT scan up to one week prior to the day of recruitment. In order to include a wide spectrum of control patients, we also recruited patients without any respiratory symptoms or suspected SARS-CoV-2 infection, and a small group of apparently healthy individuals (**Table 1**). We excluded patients with any of the following conditions: lung malignancy, rheumatic disease with shoulder or back joint involvement, acute or chronic skin disease of the chest or back, critically ill patients with mechanical ventilation, and those unable to provide informed consent.

We conducted a pilot study aimed to determine the feasability of our imaging method. This study included 55 individuals, of whom 7 were COVID-19 positive. On this pilot sample, we developed and calibrated our thermal image acquisition process, and began to develop our thermal image processing algorithms that we subsequently used in our final blinded study which comprised 101 individuals.

### 4.2 Clinical data and classification

We collected particpants’’ medical history and clinical evaluation, along with results of blood tests and CXR/CT scans. The MDclone platform was used to automatically extract multiple demographic and clinical variables from Electronic Health Records^19^.

A COVID-19 positive status was defined as having a positive SARS-CoV-2 qRT-PCR test during hospitialization or up to 10 days prior to the patient’s visit to the ER.

We classified patients according to the presence of lung injury during the time of thermal image acquisition. Lung injury was defined as any clinical or radiological evidence of pneumonia or acute respiratory distress^9,20^. Specifically, patients with any of the following conditions were defined as having lung injury: 1) any pathological findings on CXR up to 1 week before recruitment (including ground glass appearance, consolidation, and linear opacities^9^); 2) hypoxemia (≤ 93% SPO_2_ measured in room air on recruitment day); 3) significant respiratory symptoms on recruitment day (including dyspnea, respiratory rate >20, shortness of breath and/or severe cough). Control patients included individuals with no respiratory symptoms or chest abnormalities on imaging (regardless of COVID-19 status).

We acquired follow-up data on all hospitalized patients up to 3 months after imaging, including use of non-invasive oxygen therapy, use of mechanical ventilation, and death.

To compare thermal imaging scores with CXR, we manually reviewed CXRs that were acquired on the same day as thermal image acquisition (n=30), and assigned radiographic assessment of lung edema (RALE) scores^12^. The CXRs were reviewed by a certified radiologist who was blinded to the clinical status of the patients and thermal imaging scores.

### 4.3 Thermal imaging

We captured thermal images using a FLIR ONE thermal camera device (FLIR Systems, Inc. Wilsonville, OR, USA)^21^. FLIR ONE connects directly to smartphones (both IOS- and Android-operated) and utilizes the following functions: a frame rate frequency of 8.7 Hz, an object temperature range of −20°C to 120°C, and thermal sensitivity of 100 mK. The wavelength sensitivity over which the camera interpolates temperature is 8-14 µm, and the emissivity value considered appropriate for accurate human temperature readings was 0.98.

The optimal imaging procedure included images of the entire backs of the patients (**Figure 1**), who were asked to remove their top clothing, expose their backs and stand in an upright neutral position (hands down the sides of the body). Three images of the entire back were captured at a distance of ∼ 80 cm (with the back filling the entire frame of the image) (**Figure 1**). Three additional close-up images of the scapular region were also captured.

### 4.4 Thermal image processing

Thermal images of the final study cohort (n=101) were analyzed by researchers who were blinded to the patients’’ clinical status. We extracted over 100 texture and shape features of the thermal images, of which two new parameters were defined and selected for downstream analysis: one was the fractal dimension (FD) of the gradient and the other we called the “sum of extrema” (SX) in the image. The algorithm starts by reading the original thermal image (in numerical format). For the FD, the temperature matrix is first normalized to the range [0,1], followed by manual selection of the image ROI. Then a directional Sobel filter, which derivates along the vertical axis, is applied on the selected ROI. The columns of the resulting edge image are then attached one after the other, thus forming a one-dimensional vector, which is treated as a signal whose FD we wish to compute. This FD is computed using Higuchi’’s algorithm^22^ with Kmax=15.

For the SX, the temperature range in the matrix is first linearly transformed to the range [0,1], followed by manual selection of the image ROI. Then, each column of the ROI is averaged and a vector of these average values is formed. The second derivative of this vector is computed, and its absolute values from the entire vector are summed. The obtained sum provides the value of the SX, which is described below:

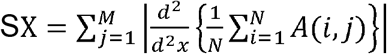

where: A(i,j) is the ROI, N is the number of rows in the ROI, and M is the number of columns in the ROI.

### 4.5 Statistical analyses and machine learning techniques

All continuous variables are displayed as mean (± standard deviation) for normally distributed variables, or median [interquartile range] for variables with nonnormal distributions. Categorical variables are displayed as counts (%) of individuals within each group.

Specific statistical tests are detailed in the figure legends. In brief, differences between values were tested by a two-tailed unpaired t-test. If values were not normally distributed, we used the non-parametric Mann-Whitney test. To assess associations among categorical variables, we used a Chi-square test. We used a one-way ANOVA with Tukey’s test for multiple comparisons to assess the significance of measurements between more than two groups. Outliers were identified by the ROUT method (Q=1%).

We used a ROC curve under the non-parametric assumption to calculate the AUC for the continuous variables “FD” and “SX” to diagnose three states: 1) COVID-19 disease status; 2) the presence of lung injury; and 3) the composite of COVID-19 status and/or lung injury. Sensitivity and specificity values were calculated for each classification as well as positive-and-negative predictive values. Spearman’s correlation test was used to assess the correlation between thermal imaging parameters and patients’’ RALE scores on CXRs, and biomarkers.

For machine learning based analysis, the dataset was split into a training and validation set. The predicted label was COVID-19 status. Three models were tested on the training set in a leave-one-out-cross-validation manner, logistic regression, k-nearest neighbors (with k=3 and with k=7) and kernel-SVM (with a radial basis kernel). The training test set included 81 data points and the validation set included 20 data points. The split was determined randomly. The experiments were repeated with the addition of age and sex variables to all three models.

Statistical analyses were performed with SPSS (IBM SPSS Statistics, version 25, IBM Corp., Armonk, NY, USA, 2016), GraphPad Prism version 8.00 (GraphPad Software, La Jolla, CA, USA), and MATLAB software (Mathworks Inc. Natick, MA, USA).

## Data Availability

The anonymized datasets generated during this study are available from the corresponding author upon reasonable request. The raw thermal images of the patients will not be available due to patient privacy regulations.

## ACKNOWLEDGMENTS

This work was performed in partial fulfillment of the requirements for a Ph.D. degree of Rafael Y. Brzezinski at the Sackler Faculty of Medicine, Tel Aviv University, Tel Aviv, Israel. We thank Vivienne York for her skillful English language editing of the manuscript. We thank Or Peretz for her help with the graphic designs of the figures, and we thank Dana Yaffe for her help with the MDClone big data platform. We thank Mrs.Lital Ben Baruch, Mr. Adam Ganim, and Mr. Erez Madaee from the School of Electrical Engineering at Afeka College, for their technical assistance in the processing of data from the thermal images.

## AUTHOR CONTRIBUTIONS

JL,RYB, NR, ZB, NNS, GR, and OH participated in study conception and design. RYB, NL, RP, AMT, OG, LS, EH, HA, IB, AL, HM, IG, SST, OR, and AI participated in patient recruitment and thermal image acquisition, as well as the collection of clinical information. OH and YZ developed the image processing algorithms and processed the thermal images. NR, OH, and RYB conducted the machine learning-based analysis.AK, EK, and EM performed the interpretation and scoring of chest X-rays.

RYB, JL, OH, NR, NL, and NNS participated in analysis and interpretation of the data. RYB, NR, JL and OH drafted the manuscript and NNS, EG, ZB, GR, and YZ helped in critical review of the manuscript. All of the authors have read and approved the final manuscript.

