## Supplementary information for "Automated processing of thermal imaging to detect COVID-19"

**Supp Figure 1: Lower back region of interest**

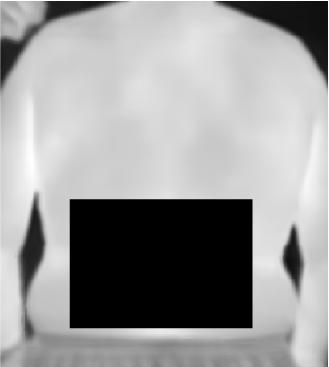

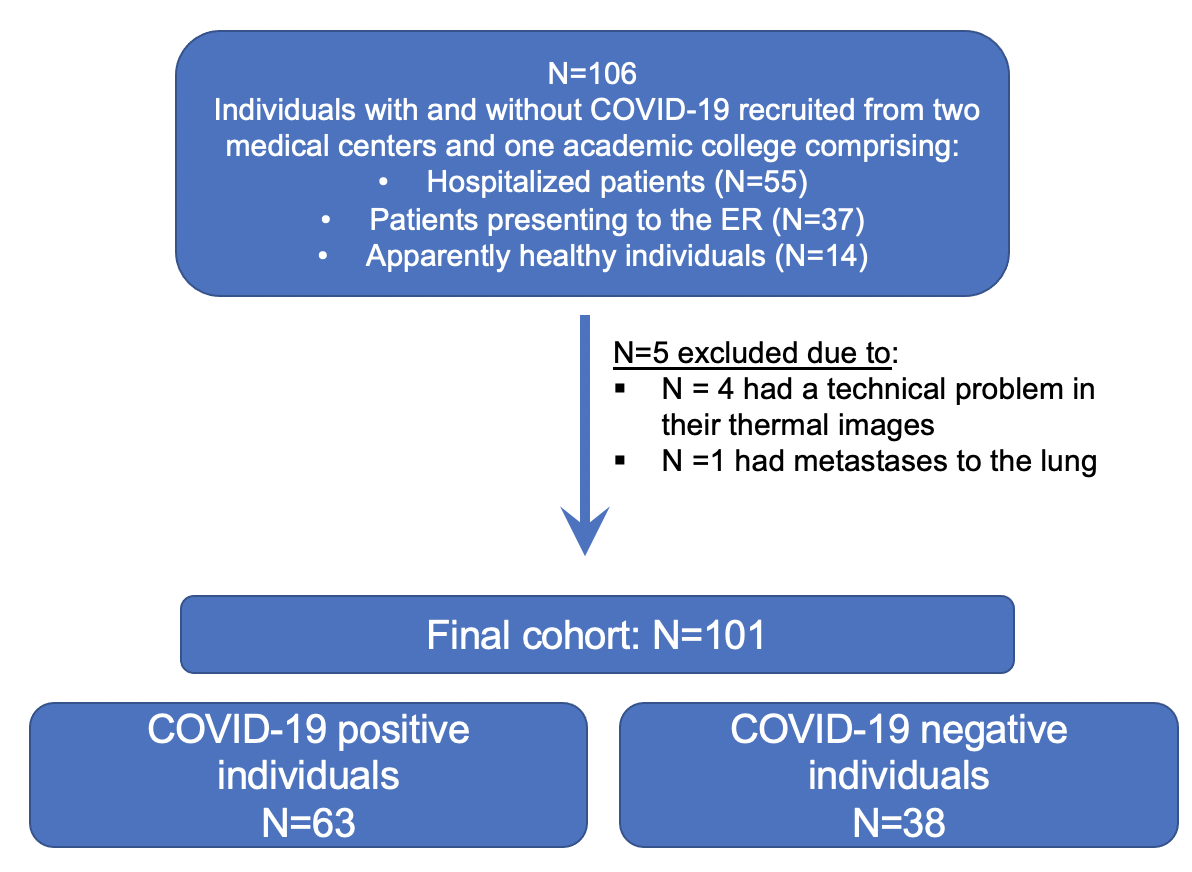
**Supp Figure 2: Research design and patient selection**

**Supp Table 1: Sensitivity and specificity values of all three machine learning models according to training and validation phases**

| Phases and variables/model | **Logistic regression** | | **k-nn (k=3)** | | **k-nn (k=7)** | | **Kernel-SVM** | |
| --- | --- | --- | --- | --- | --- | --- | --- | --- |
|  | Sensitivity | Specificity | Sensitivity | Specificity | Sensitivity | Specificity | Sensitivity | Specificity |
| **Training set:**  **FD, SX** | 82% | 84% | 78% | 66% | 80% | 63% | 88% | 59% |
| **Validation set:**  **FD, SX** | 69% | 100% | 77% | 71% | 77% | 71% | 77% | 57% |
| **Training set:**  **FD, SX, Age, Sex** | 82% | 81% | 82% | 47% | 83% | 53% | 83% | 59% |
| **Validation set:**  **FD, SX, Age, Sex** | 69% | 100% | 77% | 43% | 85% | 29% | 85% | 71% |

** FD - Fractal dimension; SX – Sum of Extrema; k-nn - k-nearest neighbors; kernel-SVM - support machine vector.*
